# An Optimization Framework to Study the Balance Between Expected Fatalities due to COVID-19 and the Reopening of U.S. Communities

**DOI:** 10.1101/2020.07.16.20152033

**Authors:** Victoria C. P. Chen, Yuan Zhou, Alireza Fallahi, Amith Viswanatha, Jingmei Yang, Yasaman Ghasemi, Nilabh. S. Ohol, Jay M. Rosenberger, Feng Liu, Xinglong Ju, Jeffrey B. Guild

**Affiliations:** Center on Stochastic Modeling, Optimization, & Statistics (COSMOS), Department of Industrial, Manufacturing, and Systems Engineering, The University of Texas at Arlington, Arlington, TX, U.S.A.; Department of Anesthesia, Massachusetts General Hospital, Harvard Medical School, Boston, MA, U.S.A.; Picower Institute for Learning and Memory, Massachusetts Institute of Technology, Cambridge, MA, U.S.A.; Department of Radiation Oncology, The University of Texas Southwestern Medical Center, Dallas, TX, U.S.A.; Department of Radiology, The University of Texas Southwestern Medical Center, Dallas, TX, U.S.A.

**Keywords:** SARS-CoV-2, COVID-19, Optimization, Trade-off, Reopening, Fatality rate

## Abstract

As communities reopen following shelter-in-place orders, they are facing two conflicting objectives. The first is to keep the COVID-19 fatality rate down. The second is to revive the U.S. economy and the livelihood of millions of Americans. In this paper, a team of researchers from the Center on Stochastic Modeling, Optimization, & Statistics (COSMOS) at the University of Texas at Arlington, in collaboration with researchers from University of Texas Southwestern Medical Center and Harvard Medical School, has formulated a computationally-efficient optimization framework, referred to as COSMOS COVID-19 Linear Programming (CC19LP), to study the delicate balance between the expected fatality rate and the level of normalcy in the community. Given the disproportionate fatality characteristics of COVID-19 among those in different age groups or with an underlying medical condition or those living with crowding, the key to the CC19LP framework is a focus on “key contacts” that separate individuals at higher risk from the rest of the population. The philosophy of CC19LP lies in maximizing protection of key contacts, so as to shield high-risk individuals from infection. Given the lack of pharmaceutical solutions, i.e., a vaccine or cure, the CC19LP framework minimizes expected fatalities by optimizing the use of non-pharmaceutical interventions, namely COVID-19 testing; personal protective equipment; and social precautions, such as distancing, hand-washing, and face coverings. Low-risk individuals that are not key contacts, including most children, are unrestricted and can choose to participate in pre-pandemic normal activities, which eliminates the need for compliance across the entire population. Consequently, the CC19LP framework demonstrates optimal strategies for protecting high-risk individuals while reopening communities.

## 1. Introduction

Widespread transmission of the novel coronavirus, SARS-CoV-2, has compromised U.S. efforts to contain the spread of the COVID-19 associated illness and subsequent fatalities. Prior guidelines to avoid the spread of viruses are based on the premise that infected individuals are most contagious after the onset of symptoms [1]. However, studies of SARS-CoV-2 have demonstrated asymptomatic and pre-symptomatic transmission [2-5], corroborated by preliminary results from antibody testing [6]. Recognized *social precautions* to prevent the spread of SARS-CoV-2 consist of 6-foot social distancing, hand-washing for at least 20 seconds with non-antibacterial soap, hand sanitizers with at least 60% alcohol content, face coverings [7], and 14-day quarantine for infected individuals [8]. Successful containment of the virus has been seen in countries with rigorous contact tracing combined with abundant COVID-19 testing capabilities, and public compliance with social precautions [9-10]. However, the U.S. faces issues with limited resources to implement contact tracing and the threat of non-compliance by lower-risk individuals [11-12].

The key question is: How do we keep the fatality rate from rising while reopening communities? Because 81% of infected individuals have mild to moderate disease [13] and may not quarantine, studies emphasize the need to isolate or shield individuals that have a higher risk of fatality [14-15]. Strategies that apply equally are in actuality unfair to those at higher risk that require interventions to equalize their risk. However, isolating high-risk groups is impractical since some need employment, require assistance, or live in environments with “crowding” and cannot isolate. In this paper, we present the COSMOS COVID-19 linear programming (CC19LP) optimization framework that enables study of the trade-off between the expected fatality rate and the normalcy of activities by “key contact” individuals in the population. CC19LP minimizes expected fatalities by optimizing the use of *non-pharmaceutical interventions*, namely social precautions, COVID-19 testing, and personal protective equipment (PPE), such as N95 respirator masks, face shields, or equivalent.

## 2. Methods

### 2.1 Population Partition

The CC19LP framework was motivated by the concept of taking care of yourself, so that you can continue to care for others [16]. In this concept, the vulnerable group only stays protected if associated caregivers remain healthy. The CC19LP framework is based on partitioning the population into three non-overlapping groups, listed below and described in Figure 1.

**Figure 1:**
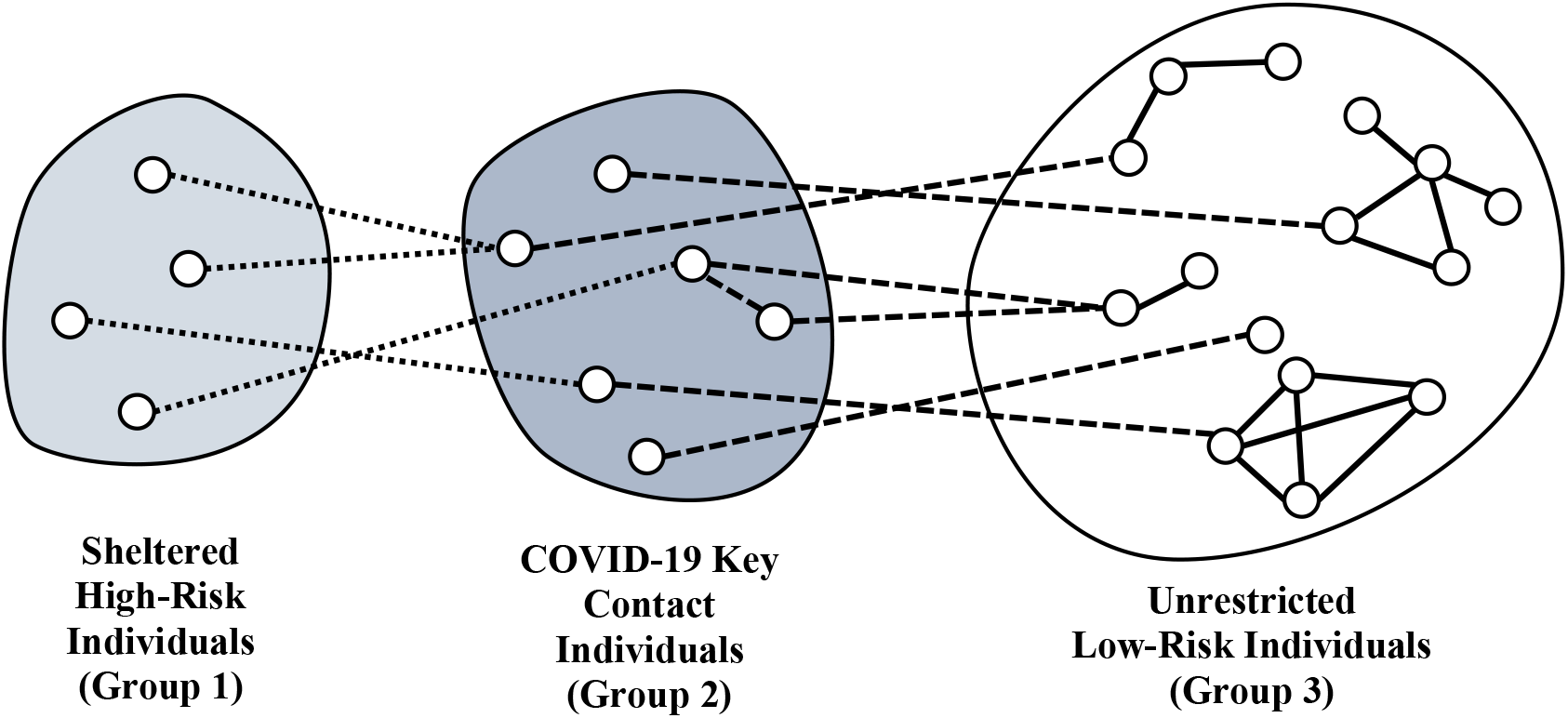
Partitioning the population into three non-overlapping groups. The role of Group 2 is to minimize transmission of the virus from Group 3 to Group 1. Dotted lines denote contacts with shelter-in-place. Dashed lines denote contacts requiring usage of social precautions, PPE, or testing. Solid lines denote unrestricted contacts.

Group 1: *Sheltered high-risk individuals* that can shelter-in-place with the assistance of key contact individuals in Group 2.

Group 2: *COVID-19 key contacts* that simultaneously have a role in protecting high-risk individuals in Group 1 and cannot avoid interaction with the rest of the population in Group 3.

Group 3: Unrestricted low-risk individuals that do not belong to Group 2.

Excluded from the population are individuals in direct contact with COVID-19 patients that are currently protected by hospital-grade PPE. Both the concepts of isolating and shielding can be incorporated in the CC19LP framework, where those that cannot avoid interaction with both the sheltered high-risk group and the unrestricted low-risk group should be classified as key contacts. High-risk children are among the sheltered high-risk group and will have a parent or guardian as a key contact. Low-risk younger children (aged 0-4) are in the unrestricted low-risk group. Low-risk children in K-12 school can be key contacts, but will likely be a small proportion of individuals because parents are more appropriate as key contacts. Recovered individuals can be key contacts or unrestricted low-risk individuals, and as information on their immunity to re-infection becomes available, their characteristics can be incorporated.

It is unrealistic and economically undesirable to maintain controls on unrestricted low-risk individuals (Group 3) for an extended period. By identifying this COVID-19 key contact population (Group 2), a large part of the population (Group 3) may return to near-normal activities while maintaining protection of high-risk individuals in Groups 1 and 2. The CC19LP framework focuses on the optimal use of non-pharmaceutical interventions specifically on key contacts (Group 2). While compliance by key contacts may be a concern, the issue of compliance is lessened because it is not applied on the entire population. The representation of social precautions is modeled as compliance in activities in the next section.

### 2.2 Activity Levels in CC19LP

In this paper, normalcy is represented on a scale of 0 to 10, where 0 mimics an environment closest to compliance with widely enforced shelter-in-place orders, and 10 mimics pre-pandemic activity. Activity levels define compliance with social precautions, where normal levels correspond to pre-pandemic activity without social precautions. Key contact children are assumed to participate in K-12 school, and key contact adults are assumed to participate in an occupation, such as employment or post-secondary school or other activities during work hours. All key contacts may additionally participate in community activities. School activities for key contact children are categorized as:

- School 1: In-person K-12 school maintaining all social precautions;
- School 2: In-person K-12 school with near-normal activities.

Occupation activities for key contact adults are categorized as:

- Work 0: Online occupation;
- Work 1: In-person occupation maintaining social precautions;
- Work 2: In-person occupation with near-normal activities.

Community activities outside of school, occupation or work hours are categorized as:

- Community 0% of normal: Maintain social precautions;
- Community 25% of normal;
- Community 50% of normal;
- Community 75% of normal;
- Community 100% of normal: Pre-pandemic normal level.

As an example, a key contact at a 25% of normal community activity level might go to occasional events (e.g., theater, restaurants) without all social precautions, but is otherwise compliant. A 50% of normal level is similar to the overall level of reported compliance among individuals aged 18-31 years prior to shelter-in-place orders [12]. A key contact at a 75% of normal community activity level might go to the gym regularly and neighborhood gatherings without social precautions, but would comply when not too inconvenient. The higher activity levels represent more normal activities, including large gatherings (e.g., church, conferences, stadium events) without social precautions.

A CC19LP optimal solution assigns key contacts to exactly one school/occupation level and exactly one community activity level in order to satisfy an overall normalcy level, and coordinates allocation of PPE and testing to minimize the expected fatality rate. The CC19LP optimization framework explicitly models key contacts that live with crowding and cannot reliably comply with social precautions. To model crowding, the community activity levels for these key contacts are forced to be at near-normal levels, while other key contacts may be restricted to lower activity levels. Constraints in CC19LP can limit the feasibility of activities, such as limiting how many can conduct their occupation completely online.

### 2.3 Expected Fatality Rate

The primary objective is to minimize the expected daily fatality rate. The calculation of the fatalities requires the following types of inputs:

- Daily number of close contacts. These contact rates increase as activity levels increase [17-19].
- Probability that a close contact is COVID-19 contagious. This uses a ratio of unknown to known cases [20] and conservatively estimates individuals to be contagious for 21 days [21].
- Transmission and fatality probabilities. For CC19LP, these were estimated based on the literature [22-23] combined with agent-based modeling.
- Effectiveness of PPE. CC19LP assumes a rate of reduction, so that cases in which PPE was not effective are represented.
- False negative rate for COVID-19 testing and quarantine probabilities. CC19LP also incorporates the uncertainty in a key contact complying with quarantine.

## 3 Computational Results

The CC19LP optimization employs linear programming, which is a computationally-fast and well-established methodology that has proven applicability across innumerable domains [e.g., 24-32]. Given the evolving nature of the pandemic, an ability to study many different input settings is an important benefit of CC19LP. There are multiple ways to define COVID-19 key contacts, and this section presents two realistic scenarios for a case study city with a total population of 400,000. Using percentages from a Gallup poll [33], this case study population has 113,250 adults classified as high-risk by age (65+) or due to underlying medical conditions. In addition, 21,950 high-risk adults are living with crowding. Based on results from an agent-based simulation of this population, the key contacts for Scenario 1 are estimated as in Table 1. There are 27,000 low-risk individuals that are key contacts, which leaves about 65% of the total population, or 259,750 unrestricted low-risk individuals, that can move towards normal activities. Key contacts with crowding are modeled separately because their activities are closer to pre-pandemic normal, so as to represent their inability to maintain social precautions.

**Table 1:** Key Contacts Composition for Scenarios 1 and 2. Individuals that live with crowding are assumed to be 15% of the population.

| COVID-19 Key Contact Type | Scenario 1 | Scenario 2 |
| --- | --- | --- |
| K-12 Children without Crowding | 1,530 | 1530 |
| K-12 Children with Crowding | 270 | 0 |
| High-risk Adult without Crowding (H2) | 28,050 | 35,100 |
| High-risk Adult with Crowding (H1) | 4,950 | 21,950 |
| Low-risk Adult without Crowding (L2) | 21,420 | 21420 |
| Low-risk Adult with Crowding (L1) | 3,780 | 0 |
| COVID-19 Key Contacts Total | 60,000 | 80,000 |

The key contact population in Scenario 2, also shown in Table 1, was augmented to include some high-risk adults without a formal occupation. While most older adults are not employed, some do not live with an appropriate low-risk key contact, and consequently must interact directly with the unrestricted low-risk group. Further, all 21,950 high-risk adults living with crowding may be unable to avoid interactions with the unrestricted low-risk group. By directly including all high-risk adults with crowding as key contacts, the defined role of low-risk key contacts is no longer needed in the crowding environment. Starting with the key contacts in Scenario 1, 4,050 low-risk key contacts with crowding were subtracted; 17,000 high-risk adults with crowding were added; and 7,050 high-risk adults without crowding were added.

### 3.1 Scenario 1 Results

The CC19LP optimization was run with a normalcy of 5 on the 0-10 scale. A total of 10,000 PPE and 150 COVID-19 tests per day were available. The optimal CC19LP solution prioritized PPE for high-risk key contacts. In particular, 100% (4950) of high-risk key contacts living with crowding were given PPE to mitigate their inability to comply with social precautions. Of the high-risk key contacts without crowding, 10% (2805) were given PPE and permitted both normal occupation and activity levels to accommodate those workers that cannot comply with social distancing, while the other 90% were required to maintain social precautions at their occupation and were limited to community activity at the lowest level. The remainder of PPE went to 5.19% (1111) of low-risk key contacts without crowding and 30% (1134) living with crowding. Of low-risk adult key contacts without PPE or tests, 96.14% (21,924) were restricted to maintain social precautions at their occupations. All 1530 K-12 key contacts without crowding and 50% (135) of those living with crowding were restricted to comply with social precautions at school. The other 50% of those in crowding were assumed to be unable to comply with social precautions at school or in community activities.

The CC19LP solution assigned all 150 tests to low-risk adult key contacts without crowding. The logical explanation for this is because tests do not prevent infection, and high-risk individuals should not risk infection. The benefit of tests is seen by key contacts quarantining when they test positive, so as not to infect sheltered high-risk individuals. Key contacts with crowding have a lower chance of complying with quarantine, so testing was not as beneficial for them. With regard to the assignment of PPE and COVID-19 tests, the CC19LP allocation consistently follows the above guidance across changes in key contact composition, changes in the magnitude of estimated probabilities and rates, and changes in the availability of PPE and COVID-19 tests. **The PPE priority order is high-risk key contacts with crowding, followed by other high-risk key contacts and low-risk adult key contacts with crowding. The priority order for COVID-19 tests is low-risk key contacts**.

With this normalcy level of 5, the expected fatality rate was estimated at 0.972 per day, equivalent to a daily U.S. fatality rate of 802. The expected fatality rate is dependent on the proportion of contagious individuals, which was estimated at 0.76%. This estimate counted confirmed COVID-19 cases in the U.S. during the last 3 weeks of April (603,793) [34] to represent known cases that could be contagious and assumed 85% would quarantine. Because U.S. testing protocol is focused on those with symptoms, estimates on the ratio of unknown to known cases can be based on the ratio of asymptomatic to symptomatic cases, which has been estimated as high as 4:1 [20].

Children without crowding are not allocated PPE or tests because they are the lowest risk group. If PPE availability is increased to 20,000, the CC19LP optimal solution shows a benefit in assigning PPE to 66.7% of these 1800 K-12 key contacts, so as to enable them to safely participate in school and community activities without social distancing. The priority, however, is still high-risk key contacts, with PPE allocated again to 100% (4950) of those living with crowding and 10.0% (2805) of those without crowding. Of the low-risk key contacts, PPE was allocated to 100% of those living with crowding (3780 K-12 and adults) and 35.7% of those without crowding (8195 K-12 and adults). The 150 tests were shifted to low-risk K-12 key contacts without crowding because low-risk adults with high activity levels are now protected by PPE. With the additional PPE, the expected fatality rate drops to 0.404 per day, equivalent to a daily U.S. fatality rate of 333.

With a desired normalcy of 9 on the 0-10 scale and the same 20,000 PPE and 150 tests per day, the expected fatality rate increases to 4.29 per day, equivalent to a daily U.S. fatality rate of 3538. However, if PPE is increased to 35,000 per day, then the expected fatality rate drops to 1.78 per day, equivalent to a daily U.S. fatality rate of 1471. This is similar to the average daily U.S. fatality rate of about 1400 from May 2020 [34], following several weeks of shelter-in-place orders. The lesson here is that a community can reopen with a higher level of normalcy among key contacts if additional PPE are available. This demonstrates the benefit of placing protective resources where they can be most effective while mitigating lower compliance with social precautions by key contacts.

### 3.2 Scenario 2 Results

For Scenario 2, the key contact population for CC19LP can be estimated based on U.S. Census data. Assuming a normalcy of 5 on the 0-10 scale and 20,000 PPE and 150 tests per day for key contacts, as in Scenario 1, the CC19LP optimal solution for Scenario 2 yields an expected fatality rate of 1.81 per day. This is 4.5 times the rate for Scenario 1, demonstrating that this larger key contact population requires more compliance with social precautions or more protective resources. As before, PPE is prioritized for high-risk key contacts at the highest activity levels. However, because of the larger population with crowding, compared to Scenario 1, the PPE supply is insufficient. The lowest normalcy attainable for Scenario 2 is 3.61 on the 0-10 scale, which yields an expected fatality rate of 1.38 per day, equivalent to a daily U.S. fatality rate of 1140. Figure 2 summarizes the CC19LP optimal solutions, showing the best use of non-pharmaceutical interventions on key contacts. Comparing the solutions for the two overall normalcy levels, 5 and 3.61, the allocation of PPE and testing, indicated by pulled-out slices, is identical. To achieve the lower normalcy of 3.61, the primary reduction in activity levels is seen with low-risk key contacts (K-12 and L1), where community activity is reduced to the lowest level that requires compliance with all social precautions.

**Figure 2:**
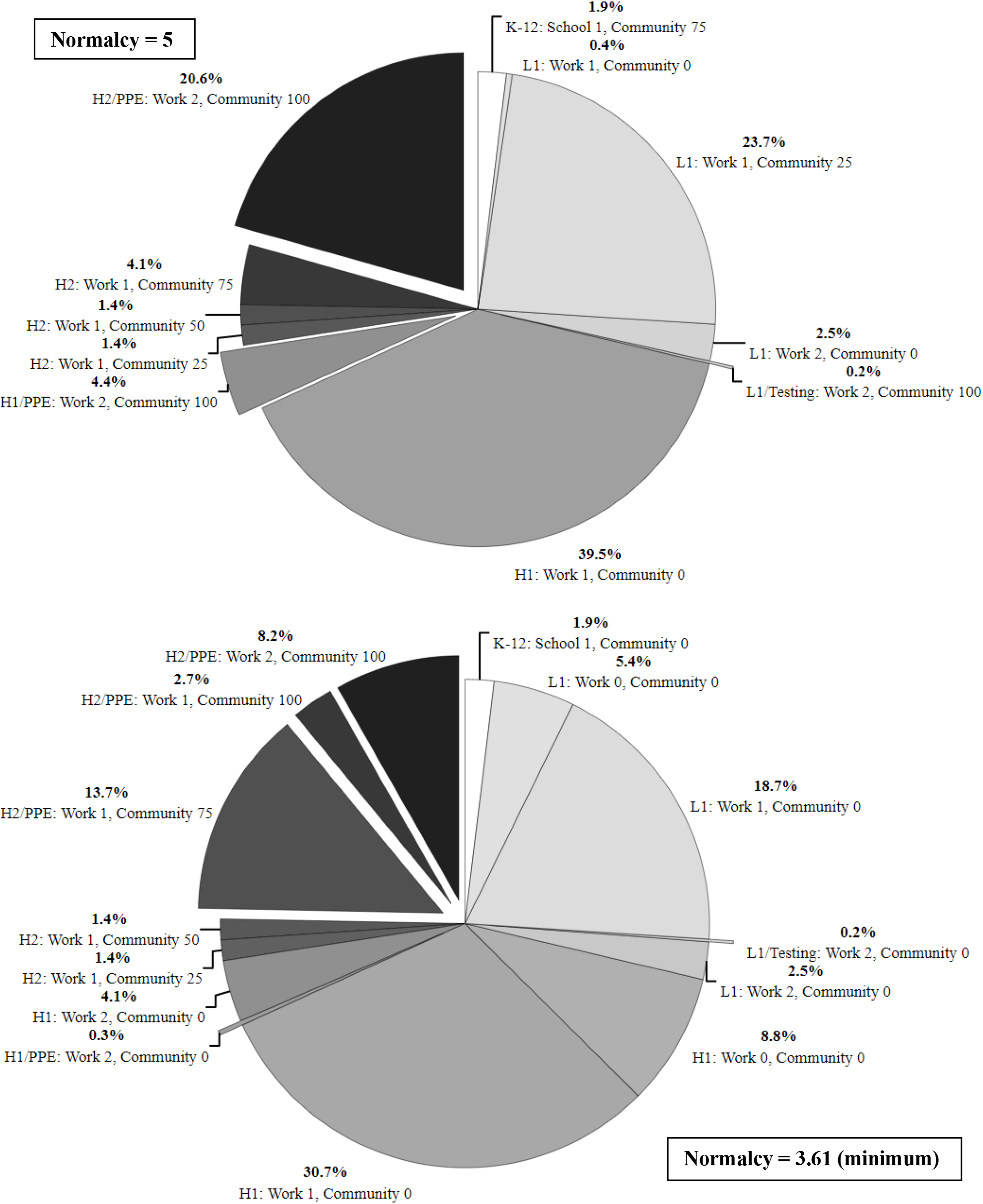
CC19LP solutions for Scenario 2 key contacts with normalcy 5 (top) and 3.61 (bottom). White = K-12 children. Light Grey = Low-risk adults (L1). Medium Grey = High-risk without crowding (H1). Dark Grey = High-risk living with crowding (H2). See Section 2.2 for notation on activity levels. PPE and COVID-19 testing interventions are indicated by pulled-out pie slices.

To represent the U.S. population during shelter-in-place orders, a baseline CC19LP run was executed at a 3.61 normalcy. Only 5652 PPE was available per day, specifically for those in healthcare that cannot avoid close contact with high-risk patients. Testing availability was 138 per day, obtained by taking half the average daily testing in the U.S. during the week of April 26 [34] and scaling it to a population of 400,000. Table 2 shows the expected fatality rates. Taking into consideration that reported fatalities could be only 65% of actual [35] and much of the U.S. was under shelter-in-place orders during April, a ratio around 2:1 unknown to known cases could be a reasonable match to the average U.S. fatality rate from May 2020. If the low-risk group approaches herd immunity, then lower ratios could be used. CC19LP optimal interventions are not affected by the ratio of unknown to known cases.

**Table 2:** Expected fatality rate estimates for Scenario 2 baseline run under different assumptions, using the equivalent daily U.S. fatality rate.

| Quarantine Compliance | Ratio of Unknown to Known Cases |  |  |  |  |
| --- | --- | --- | --- | --- | --- |
|  | 5:1 | 4:1 | 3:1 | 2:1 | 1:1 |
| 85% | 4517 | 3640 | 2763 | 1886 | 1009 |
| 100% | 4385 | 3508 | 2631 | 1754 | 877 |

## 4 Discussion

Consider a family with two low-risk K-12 children, two low-risk working parents, and two high-risk grandparents. While the grandparents can mostly isolate, they still need sustenance and assistive care. If all four low-risk family members are required close contact with the grandparents, then all four would be key contacts. However, it is more efficient if only one family member, perhaps the son or daughter of the grandparents, is the key contact, and the other three low-risk individuals are cautioned to stay distant. In a retirement home, all staff members that interact with residents should be key contacts, and in-person visitation is not possible, unless the visitor is also a key contact. While high-risk key contacts would be expected to maintain social precautions to protect themselves, a characteristic of low-risk key contacts is a responsibility towards the sheltered high-risk group. Examples include the responsibility nurses have towards patients and individuals have towards loved ones.

The U.S. population insists upon freedom and equality. This system of COVID-19 key contacts seeks to equalize fatality risk by directing non-pharmaceutical interventions to reduce infection risk among groups with higher fatality rates while simultaneously enabling more freedom among the unrestricted low-risk group and facilitating the right to employment for key contacts. It is up to policy-makers to create processes to identify COVID-19 key contacts in the U.S. population and promote appropriate protective resources. Policy-makers can create structure that specifies eligibility and benefits. High-risk individuals can be identified by age or by medical experts and can be classified as key contacts if they cannot avoid interaction with the unrestricted low-risk group. Unrestricted individuals freely choose their activity level. Sheltered high-risk individuals should be able to identify their associated key contacts. Further, if an occupation requires close contact with high-risk individuals, such as healthcare departments, law enforcement, and fire departments, then the employer can identify employees as key contacts. For key contacts identified via employment, the requirements and benefits can be defined by the employer. For others identified by their home environment, serving as a key contact should be a choice. Benefits to encourage participation can be defined by policies, such financial resources to maintain PPE, discounts or free delivery services for shopping, and reserved sections or time periods at restaurants and venues. Since the unrestricted low-risk group will be a majority of the population, the majority of business can correspondingly benefit from our COVID-19 key contact structure.

## 5 Concluding Remarks

The CC19LP framework is a proactive planning approach to protect high-risk individuals and enable a controlled return towards normalcy. CC19LP has demonstrated that more resources targeted for key contacts can reduce the expected fatality rate without requiring restrictions on the entire population. As seen in the optimal solutions from CC19LP, PPE and COVID-19 testing are critical for key contacts that cannot avoid a high level of close contact activity with high-risk individuals. Consistent implementation of PPE and testing requires daily use by key contacts that have high activity levels. CC19LP optimal solutions focus on prevention of transmission to high-risk individuals, in particular, assuming the existence of *unknown* cases. This in contrast to current U.S. policies for PPE usage and COVID-19 testing, which focus on *known* cases and *known* symptoms. The reliance on known cases becomes less effective as infections rise among the unrestricted low-risk population, and continuing restrictions on this low-risk group is not in the best interest of the U.S. economy.

To keep the expected fatality rate down as the infection rate rises among the unrestricted low-risk population, the normalcy level for key contacts without crowding may lean conservatively towards maintaining social precautions. Governments, businesses, and organizations should create special concessions and financial support, if necessary, for key contacts, so as to preserve the right to employment while accommodating recommended social precautions, PPE, or testing, **regardless of socio-economic status**. Some businesses are currently implementing concessions, such as protective barriers for employees, special grocery store hours for the high-risk population, restaurant take-out with minimal contact, and personnel scheduling [35-38]. According to the Gallup poll [33], about 35% of the adult U.S. population is high-risk for COVID-19 complications, and 4.5% is severely high-risk. Out of a U.S. population of about 330 million, 257 million are 18 years and older, which means there are 90 million at high-risk and 11 million severely high-risk Americans. The U.S. can still choose to protect these individuals until a vaccine or cure is available. Further, once available, priority for vaccinations could follow CC19LP guidance. Specifically, those key contacts to which CC19LP allocates PPE and testing should be prioritized for vaccination. This will enable continued minimization of the expected fatality rate until the larger population can be vaccinated. An online CC19LP tool is available for public use via the COSMOS website projects page (https://cosmos.uta.edu/projects/covid-19/).

## Data Availability

An online tool for the optimization has been developed by the authors and is publicly available. Data on COVID-19 cases and fatalities were obtained from a publicly available website. Data for the optimization were generated via publicly available Census data and a Gallup poll.

https://cosmos.uta.edu/projects/covid-19/

https://ourworldindata.org/coronavirus

https://www.census.gov/data.html

https://news.gallup.com/poll/304643/million-severe-risk-infected-covid.aspx

## Acknowledgment

The authors thank Norman H. Edelman, M.D. for sharing his perspective on the COVID-19 pandemic. Dr. Edelman is Professor of Medicine; Core Faculty, Program in Public Health. Stony Brook University.

